# The Sequence Effect Worsens over Time in Parkinson’s disease and Responds to Open and Closed-Loop Subthalamic Nucleus Deep Brain Stimulation

**DOI:** 10.1101/2022.03.07.22270923

**Authors:** Yasmine Kehnemouyi, Matthew Petrucci, Kevin Wilkins, Helen Bronte-Stewart

## Abstract

**Background:** The sequence effect is the progressive deterioration in speech, limb movement, and gait that leads to an inability to communicate, manipulate objects or walk without freezing of gait. Many studies have demonstrated a lack of improvement of the sequence effect from dopaminergic medication, however few studies have studied the metric over time or investigated the effect of open and closed-loop deep brain stimulation in people with PD.

**Objective:** To investigate whether the sequence effect worsens over time and/or is improved on clinical (open-loop) and closed-loop deep brain stimulation (DBS).

**Methods:** Twenty-one people with PD with bilateral STN DBS performed thirty seconds of instrumented repetitive wrist flexion extension and the MDS-UPDRS III off therapy, prior to activation of DBS and every six months for up to three years. A sub-cohort of ten people performed the task during randomized presentations of different intensities of STN DBS.

**Results:** The sequence effect was highly correlated with the overall MDS-UPDRS III score and the bradykinesia sub-score and worsened over three years. Increasing intensities of STN open-loop DBS improved the sequence effect and one subject demonstrated improvement on both open-loop and even further improvement on closed-loop DBS.

**Conclusions:** Sequence effect in limb bradykinesia worsened over time off therapy due to disease progression but improved on open and closed-loop DBS. These results demonstrate that DBS is a useful treatment of the debilitating effects of the sequence effect in limb bradykinesia and that closed-loop DBS may offer added improvement.

## Introduction

The sequence effect is the progressive deterioration in ongoing movement that is not related to peripheral muscle fatigue [1] and it appears to be specific to Parkinson’s disease (PD) [2,3]. Although initially thought to be a feature of more advanced stage PD, the sequence effect in speech and limb movement has been documented in very early stages of PD in drug naïve people [4–7]. The sequence effect is one of the most debilitating features of PD and a significant source of morbidity: the progressive shortening of step length has been shown to cause freezing of gait (FOG), which frequently results in falls [8–12]. It has been stated that FOG results from the superimposition of the sequence effect on gait hypokinesia and that FOG will not occur in its absence [9–11]. The sequence effect in limb bradykinesia is evident in both the amplitude and frequency of ongoing movements and impairs the ability to write, use a computer keyboard and manipulate objects such as tools or buttons [3,13–18]. The sequence effect in speech in Parkinson’s disease causes reduced intelligibility, articulatory imprecision, and altered rates of speech, which affects up to 90% of people with PD, worsens with disease progression, and causes significant morbidity [19,20].

Although different components of bradykinesia can be treated with levodopa and/or deep brain stimulation (DBS), a critical unmet need for improving the lives of people with PD is that the sequence effect does not respond to dopaminergic medication [1,3,4,20–22]. Few studies have specifically studied the effect of open-loop deep brain stimulation (DBS) on the sequence effect [18,23]. However, there is some preliminary evidence that closed-loop deep brain stimulation may be equal or superior to open-loop DBS in improving the sequence effect [24,25]. Thus, further investigation into the use of open-loop and closed-loop DBS is necessary.

Our goals in this study were: (1) to investigate whether an objective measure of the sequence effect, measured using the best fit decay of angular velocity, progressed over three years in the off therapy state, (2) to explore whether the sequence effect of limb bradykinesia improved during open-loop (continuous) subthalamic nucleus (STN) DBS and whether there was a ‘dose” or DBS intensity dependence, and (3) to provide pilot data that demonstrated improvement in the sequence effect during neural closed-loop (adaptive) STN DBS.

## Materials and Methods

### Human subjects

Twenty-one individuals (5 female) with clinically established Parkinson’s disease underwent bilateral implantation of DBS leads (model 3389, Medtronic PLC) in the STN. The two leads were connected to the implanted investigative neurostimulator (Activa^®^ PC+S, Medtronic PLC, FDA Investigational Device Exemption (IDE) approved, clinical trials NCT02384421 and NCT01990313). The preoperative selection criteria and surgical technique have been previously described [26,27]. One participant in the cohort (male) was later re-implanted with another investigative neurostimulator (Summit™ RC+S, IDE approved, clinical trial NCT04043403) and completed the closed-loop study. All participants gave written consent to participate in the study, which was approved by the Food and Drug Administration (FDA) and the Stanford University School of Medicine Institutional Review Board (IRB).

### Experimental Protocol

#### Off Stimulation Longitudinal Testing

Experiments were conducted before the initial activation of the DBS system at initial programming (IP), which took place 1 month after implantation of the DBS leads, off all therapy after 6 months, and then 1, 2, and 3 years after the IP visit in 6-month intervals. All experimental testing was done in the off-medication state, which entailed the withdrawal of long-acting dopamine agonists for 48 hours, dopamine agonists and controlled release carbidopa/levodopa for 24 hours, and short acting medication for 12 hours prior to the study visit. At the follow-up visits, stimulation was turned off for 60-75 minutes. The participant then performed a repetitive wrist-flexion extension (rWFE) task, which we have previously validated as a measure of bradykinesia in Parkinson’s disease [28–31]. Participants were instructed to remain seated and as still as possible with their eyes open, and after a “Go” command, to flex and extend the hand at the wrist joint as quickly as possible and to stop only when instructed; the forearm was flexed so that the elbow was angled at 90°. The movement was self-paced and lasted 30 seconds. MDS-Unified Parkinson’s Disease Rating Scale – motor subscale (MDS-UPDRS III) was performed by a certified rater pre-operatively (on and off medication), off medication at IP, and then off all therapy at all follow-up visits after DBS had been turned off for at least 60 minutes.

#### Titrations

A sub-cohort of 10 individuals (3 female) completed a stimulation amplitude titrations experiment off medication. Participants performed five trials of the rWFE task. Each trial was performed during randomized presentations of STN DBS at 0% (no DBS), 25%, 50%, 75%, and 100% of V_max_. V_max_ represented the clinically equivalent DBS intensity using a single active electrode, with which neurostimulation improved bradykinesia to a similar degree to that observed when using the clinical DBS intensity delivered through one or multiple electrodes. Each participant performed one round of the entire experiment at the designated visit.

#### Closed-Loop DBS

Data were collected during the rWFE task in three stimulation conditions: off stimulation, clinical open-loop stimulation (olDBS), and neural closed-loop stimulation (NclDBS). The NclDBS condition used the beta burst driven adaptive closed-loop algorithm [32] with the addition of a stimulation delay period control parameter (i.e., the minimum duration of time paused after one stimulation decision before the next allowable stimulation decision). The stimulation delay period was varied from 800-2000 ms between stimulation decisions. The stimulation parameters for olDBS were (LSTN: C+,1-,2-at 4.8 mA; RSTN: C+, 8-,9-at 5.2 mA; 140 Hz) and NclDBS (LSTN: C+, 1-, 3.2-4.0 mA; RSTN: C+, 9-, 3.4-4.2 mA; 140 Hz).

### Data Acquisition and Analysis

#### Kinematic Data Acquisition

Movement was measured using solid-state gyroscopic wearable sensors (sampled at 1 kHz) attached to the dorsum of each hand and each foot (Motus Bioengineering, Inc, Benicia, CA), a triaxial accelerometer on the forehead (ADXL335, Analog Devices, Norewood, MA.), and with synchronized video recordings from a USB web camera (C930e, Logitech, Lausanne, Switzerland). Sampling rates for the gyroscope, accelerometer, and video data were 1 kHz, 1 kHz, and 30 frames per second, respectively.

#### Kinematic Data Analysis

For each movement epoch, an automated algorithm was used to quantify the sequence effect. The angular velocity data was low-pass filtered in MATLAB using zero-lag 4^th^ order Butterworth filters with a 4 Hz cutoff frequency at the above sampling frequency. Peaks for analysis were chosen based on the maximum angular velocity between each zero crossing for each cycle of flexion-extension. For traces with excess tremor that required extra smoothing to find accurate zero crossings, the angular velocity data was low-pass filtered again using zero-lag 4^th^ order Butterworth filters starting with a 2.5 Hz cutoff frequency, and if subsequent filtering was required the data was re-filtered using a cutoff frequency that decreased by 0.5 Hz until no further filtering was required or the cutoff frequency decreased to 0 Hz, at which case the trace was not usable as zero crossings could not be accurately detected. This filtered trace was only used to identify the zero crossings, which were then applied to the peak detection on the original angular velocity data.

Since some trials displayed multiple epochs of sequence effect in which the participant was able to reset following initial decrements in angular velocity, an automated algorithm was used to determine if the trial should be broken up into one or multiple epochs. First, a 3-point moving average was calculated on the angular velocity peaks to better dynamically visualize and model the trend in behavior as well as filter out periodic fluctuations and noise. To further protect the sequence effect models against overfitting from sudden behavioral fluctuations, the percent change was calculated on the moving average of peaks in two ways: between the current peak and the subsequent peak (denoted as PC1) as well as between the current peak and the next 2 peaks (denoted as PC2). A negative percent change represented a smaller angular velocity from previous, typically seen during the sequence effect epoch, and a positive percent change represented a larger angular velocity from previous, which if large enough represented the end of a sequence effect epoch. Upon inspection across the cohort’s rWFE traces, a threshold of 20% was empirically derived, where once the percent change in angular velocity crossed this threshold, a sequence effect epoch had been completed.

Following epoching, an exponential curve was fit to the first epoch of decay in a trace using the following criteria: for those where the maximum angular velocity is greater than 100 degrees/second and there are at least 10 full cycles of rWFE present, the initial point of the fitted exponential was chosen as the maximum of the first 10 peaks. For traces where the maximum angular velocity was less than 100 degrees/second and there were at least 5 full cycles of rWFE present, the initial point was chosen as the maximum of the first 5 peaks. For traces that fit neither criterion, the initial point was chosen as the first peak. The algorithm starts a new epoch when either the PC1 or PC2 trace crossed the 20% threshold and the current peak is at least 40% of the first epoch’s maximum peak (representing an accurate pick up in behavior). When this is true, the first epoch will end with the last point with negative percent change prior to crossing the 20% threshold and the second epoch will begin to be fit with an exponential function, and this process repeats until the entire trace’s sequence effect epochs have been fit with exponential curves.

The sequence effect behavior in each epoch was modeled using the exponential decay function equation:

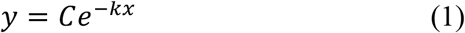

Where *y* = movement amplitude, *C* = the model intercept, *x* = time, and *k* = the slope of the decay. The slope of the decay was normalized by the initial movement velocity, *A*, to take into account the speed of the movement. A natural log was then used to normalize the distribution of the data:

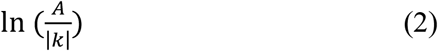

Finally, to express the sequence effect metric as a percentage where a higher number was indicative of greater (i.e., worse) sequence effect, the inverse of equation 2 was used and multiplied by 100:

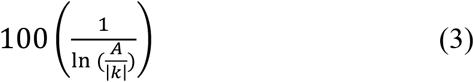

For cases where the epoch is overall increasing and an exponential growth curve was more suitable for modeling the behavior, the epoch was modeled using equation 1 with a positive exponent, and the sequence effect metric was still calculated with equation 3, but in this case the *A* coefficient in the metric was taken as the maximum angular velocity that the participant obtained in the epoch and *k* represented the slope of growth. Periods during the epoch in which tremor superseded the movement were also removed from analysis. Though this proposed modeling algorithm ran automatically, for cases where the outputted sequence effect model was visually not representative of the data, manual adjustments were made, such as shifting the starting point or increasing the percentage threshold. Finally, though in some traces there were multiple epochs of sequence effect fit by the algorithm, to avoid confounding the sequence effect metric only the first decay epoch was denoted as the primary epoch of sequence effect for analysis.

### Statistical Analysis

Statistics were computed using MATLAB (version 9.9, The MathWorks Inc. Natick, MA, USA). Pearson correlations were used to assess associations between the sequence effect during rWFE and both total MDS-UPDRS III scores and the bradykinesia sub-score. A linear mixed effects regression model was performed to analyze the effect of time (in months) on sequence effect. In this case, the sequence effect metric was included as the dependent variable and visit month was used as a fixed effect with subject as a random intercept and time as a random slope. For analysis of the titrations results, Kolmogorov-Smirnov tests and theoretical-sample quantile plots were used to assess the normality of the distribution of sequence effect at each stimulation condition. Based on the titrations experiment results, a repeated measures ANOVA was used to compare the effect of stimulation level on sequence effect across all STNs at the five stimulation conditions. Paired t-tests were used *post hoc* to compare the difference in sequence effect between the 0–25%, 0– 50%, 0–75%, and 0–100% V_max_ DBS conditions. *P*-values for *post hoc* tests were corrected for multiple comparisons (N=4) using a Bonferroni correction. A corrected p value < 0.05 was considered significant.

## Results

For the longitudinal off-stimulation study, kinematic data from 42 hands from 21 well-characterized individuals with Parkinson’s disease were included in the analysis. The average number of visits was 5.2 ± 1.6 over a span of up to 36 months (average maximum visit month=27.7 ± 10.1, Table 1). Preoperative OFF and ON medication UPDRS III scores were 43.9 ± 11.5 and 20 ± 9.7 respectively. Note, preoperative UPDRS-III scores were reported for the earliest 5 participants, while the revised MDS-UPDRS-III was utilized for the remaining 16 participants. For the titrations experiment, kinematic data from 16 hands from 10 well-characterized individuals with Parkinson’s disease were included in the analysis. This experiment was conducted 2.5 ± 0.6 years after the participant’s initial programming visit (Table 2). The mean age of the participants was 55.3 ± 9.0 years, and mean disease duration was 8.9 ± 3.1 years. Preoperative OFF and ON medication UPDRS-III scores were 41.8 ± 11.6 and 18.6 ± 7.6, respectively. OFF medication UPDRS-III scores, off and on STN DBS were 39.6 ± 14.6 and 10.7 ± 7.2, respectively, at the time of the experiment.

**Table 1.** Participant Demographics

| Participant | Sex | Disease duration at IP | Pre-Op UPDRS III (off meds) | Pre-Op UPDRS III (on meds) | IP MDS-UPDRS III | # of Visits | Max Visit Month | Time of Titration Experiment |
| --- | --- | --- | --- | --- | --- | --- | --- | --- |
| 1 | M | 10 | 54 | 41 | 60 | 4 | 24 | - |
| 2 | M | 5 | 24 | 8 | 20 | 7 | 36 | 3.3 |
| 3 | M | 3 | 35 | 26 | 36 | 6 | 36 | 3.3* |
| 4 | M | 5 | 29 | 16 | 38 | 4 | 36 | 3 |
| 5 | M | 3 | 39 | 23 | 43 | 7 | 36 | 2.8 |
| 6 | M | 4 | 58 | 12 | 51 | 5 | 30 | 2.8* |
| 7 | M | 10 | 41 | 28 | 43 | 2 | 6 | - |
| 8 | M | 7 | 52 | 29 | 61 | 6 | 36 | 2.6* |
| 9 | M | 9 | 47 | 23 | 45 | 6 | 30 | - |
| 10 | F | 10 | 33 | 6 | 42 | 6 | 36 | - |
| 11 | M | 11 | 56 | 20 | 60 | 2 | 6 | - |
| 12 | M | 2 | 59 | 22 | 70 | 6 | 30 | 1.9 |
| 13 | M | 6 | 62 | 23 | 56 | 3 | 12 | - |
| 14 | F | 6 | 44 | 25 | 12 | 7 | 36 | - |
| 15 | F | 9 | 50 | 26 | 51 | 6 | 30 | 1.9* |
| 16 | M | 14 | 42 | 24 | 44 | 4 | 18 | - |
| 17 | F | 12 | 34 | 6 | 22 | 7 | 36 | 1.8 |
| 18 | M | 7 | 51 | 10 | 33 | 5 | 30 | - |
| 19 | F | 11 | 38 | 18 | 34 | 7 | 36 | 1.6 |
| 20 | M | 9 | 11 | 7 | 22 | 4 | 18 | - |
| 21 | M | 10 | 38 | 31 | 48 | 5 | 24 | - |
| Average ± SD | | 8.0 ± 3.3 | 42.7 ± 12.7 | 20.2 ± 9.3 | 42.4 ± 15.1 | 5.2 ± 9.3 | 27.7 ± 10.1 | 2.5 ± 0.6 |

IP, initial programming; UPDRS, Unified Parkinson's disease rating scale; MDS, Movement Disorder Society; \* Indicates only 1 hand tested

**Table 2:** Sequence effect values off stimulation (OFF), on open-loop DBS (olDBS), and neural closed-loop (NclDBS) with varying stimulation delay values.

| **Right Hand** |  |  | **Left Hand** |  |  |
| --- | --- | --- | --- | --- | --- |
| **Condition** | **Delay (ms)** | **Sequence Effect** | **Condition** | **Delay (ms)** | **Sequence Effect** |
| OFF | - | 13.23% | OFF | - | 13.55% |
| olDBS | - | 10.55% | olDBS | - | 10.03% |
| NclDBS | 800 | 9.52% | NclDBS | 800 | 9.70% |
| NclDBS | 1000 | 9.74% | NclDBS | 1000 | 9.84% |
| NclDBS | 1200 | 9.46% | NclDBS | 1200 | 10.51% |
| NclDBS | 1500 | 9.03% | NclDBS | 1500 | 10.59% |
| NclDBS | 2000 | 9.82% | NclDBS | 2000 | 9.46% |

### Quantification of sequence effect epochs

Figure 1 presents the output of the algorithm, which fits the rWFE angular velocity trace with a characteristic exponential decay curve. Figure 1A displays the behavior of an individual with minimal sequence effect, represented by a relatively smaller sequence effect metric. Figure 1B shows an individual with more visible sequence effect whose dynamics were modeled by one exponential decay curve. Figure 1C portrays an example of an individual who experienced 2 epochs of sequence effect as modeled using 2 separate exponential decay curves by the algorithm. At the initial visit of the longitudinal off-stimulation study, 25 hands showed only one epoch of sequence effect and 17 exhibited multiple epochs. Across the 21 participants throughout the 3 years of repeat visits, 134 trials showed only one epoch of sequence effect and 78 trials showed multiple epochs of sequence effect.

**Figure 1.**
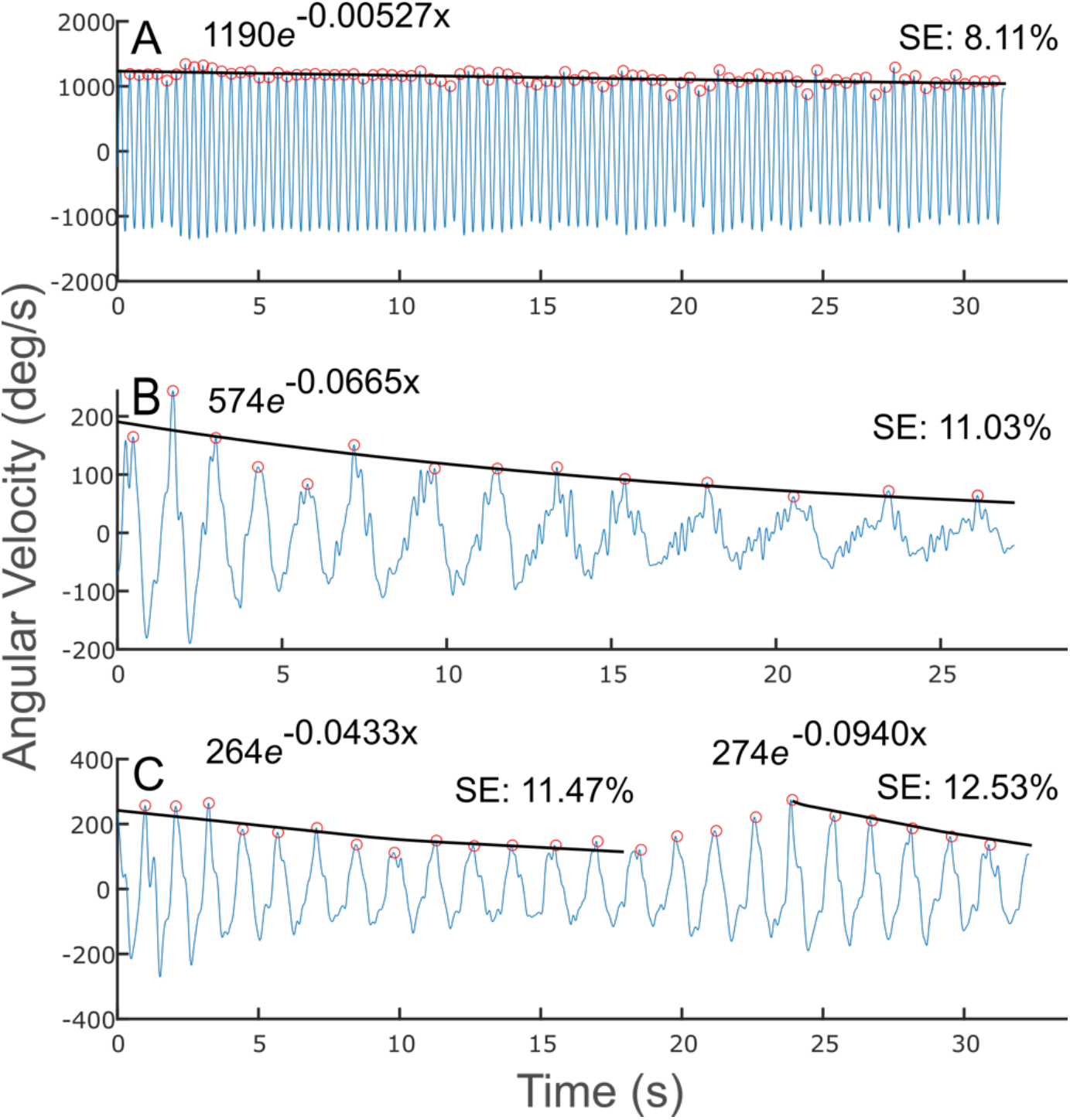
Quantification of sequence effect on example WFE traces. (A) WFE trace with minimal sequence effect. (B) WFE with one epoch of substantial sequence effect for the entire trace. (C) WFE split into two epochs based on reemergence of performance around 25 seconds. Fitted exponential line is shown in black. Detected peaks shown by orange open circles. Exponential fit function and subsequent sequence effect (SE) metric shown above each trace.

### Sequence effect is related to overall motor impairment and bradykinesia

Figure 2 shows the comparison between the measured sequence effect (n=42 hands) versus total MDS-UPDRS III score (Fig. 2A) and the bradykinesia sub-score for the tested side (Fig. 2B) in participants in the longitudinal study after performing the rWFE task at their initial programming visit. Higher (i.e., worse) sequence effect was associated with greater total impairment (r = 0.59, *p* = 4.81e-5) and bradykinesia (r = 0.59, *p* = 4.09e-5).

**Figure 2.**
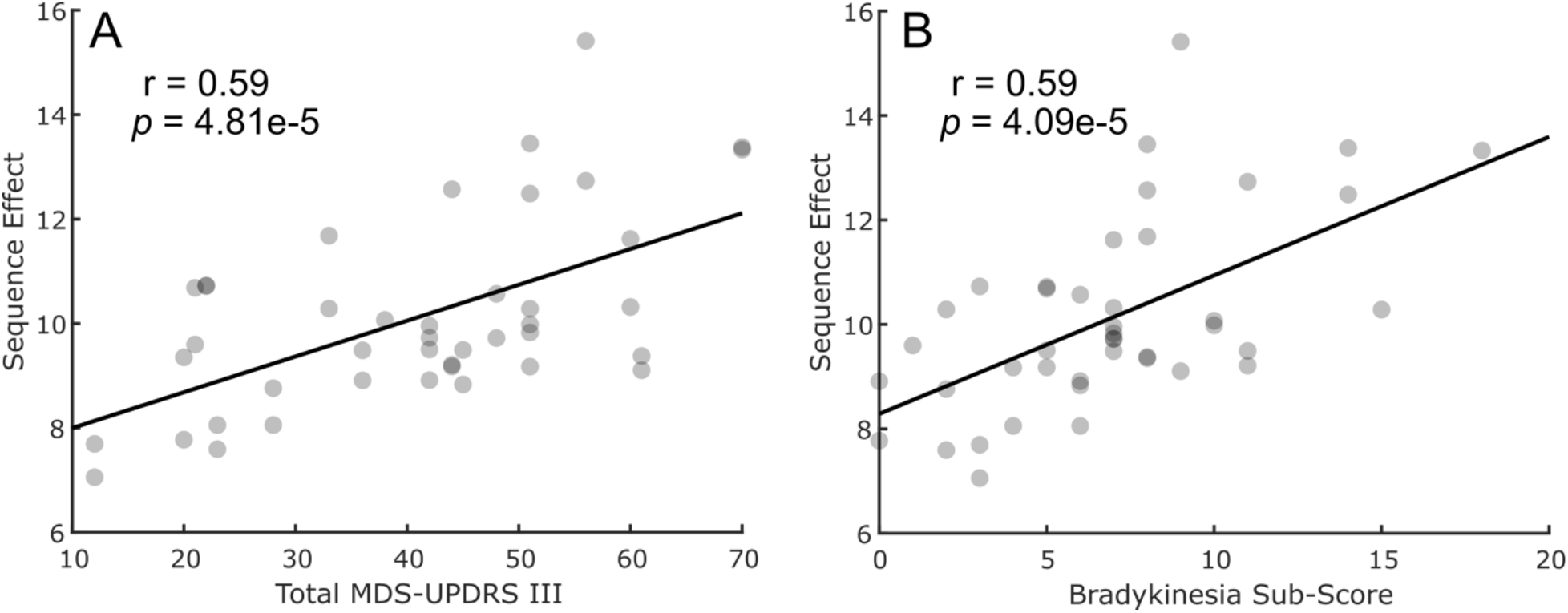
Scatter plot between sequence effect during rWFE and (A) total MDS-UPDRS III scores and (B) bradykinesia sub-scores.

### Sequence effect worsens over time

Figure 3 shows the sequence effect across the longitudinal group (n=42 hands) after performing the rWFE task off therapy, at up to 7 timepoints out to 3 years after the initial programming visit. Sequence effect significantly increased (i.e., worsened) over time, as measured off therapy (β = 0.0453 (95% CI: 0.0328 to 0.0578), t = 3.62, *p* = 0.00037).

**Figure 3.**
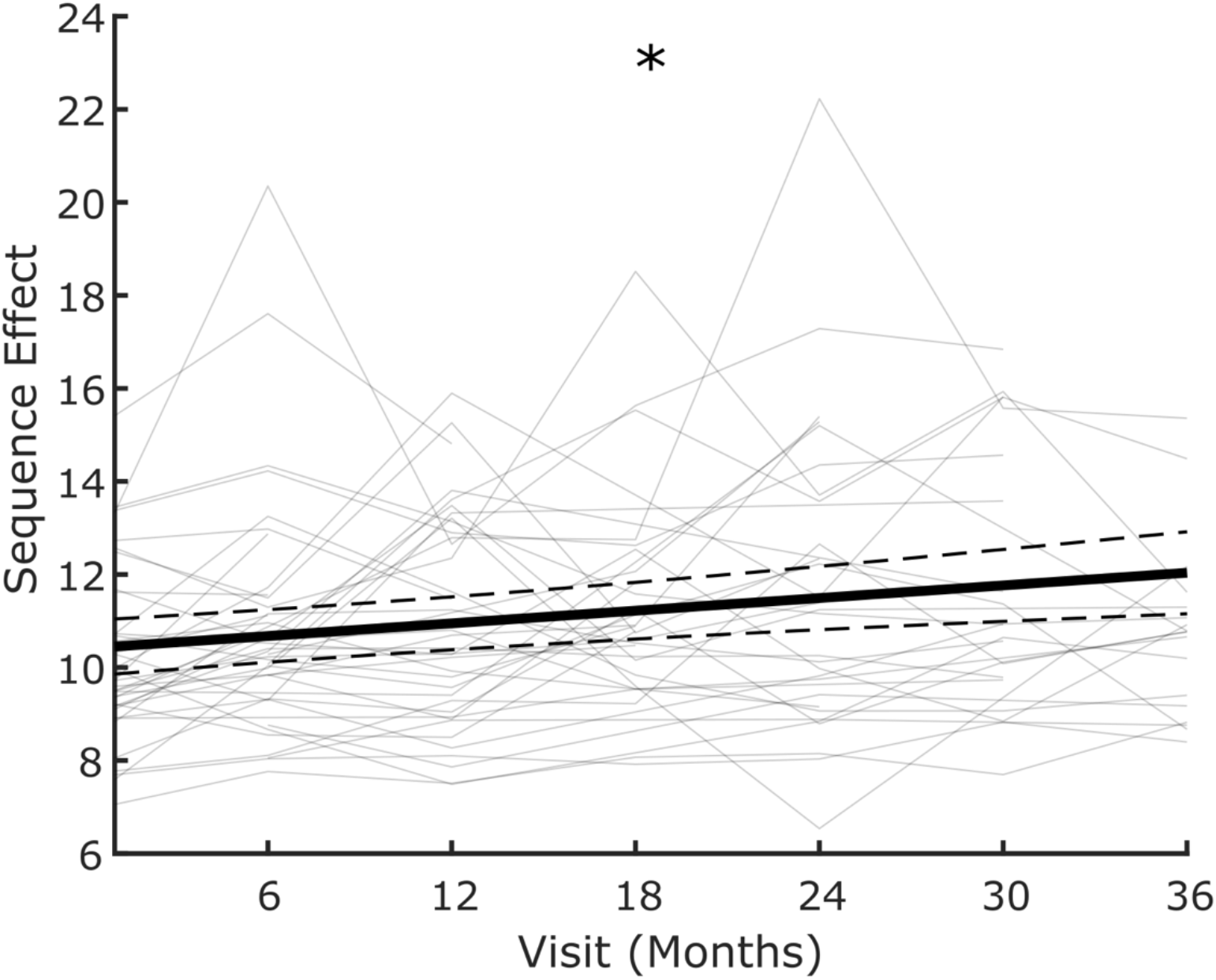
Sequence effect worsens over time off therapy. Average slope of change over time (thick black line) with individual data overlaid as line plots (light gray) of sequence effect. * indicates significant change over time. Dashed lines represent the 95% confidence interval of the slope estimate.

### STN DBS improves the sequence effect

Figure 4 shows the box plot of sequence effect across in the titration experiment (n=16 hands) after performing the rWFE task during the randomized presentations of STN DBS at 0%, 25%, 50%, 75%, and 100% of V_max_. Across the group, increasing intensities of STN DBS during rWFE were associated with a decrease (i.e., improvement) in sequence effect [*F*(4,60) = 3.01; ; *P* = 0.0233]. *Post hoc* paired t-tests tests showed significant decreases in sequence effect during the 75% and 100% V_max_ stimulation conditions: 0% and 75% V_max_ (t(15) = 3.14, *p* = 0.027), 0% and 100% V_max_ (t(15) = 3.80, *p* = 0.0068). Therefore, DBS progressively reduced sequence effect, with significant differences occurring at 75% and 100% V_max_.

**Figure 4.**
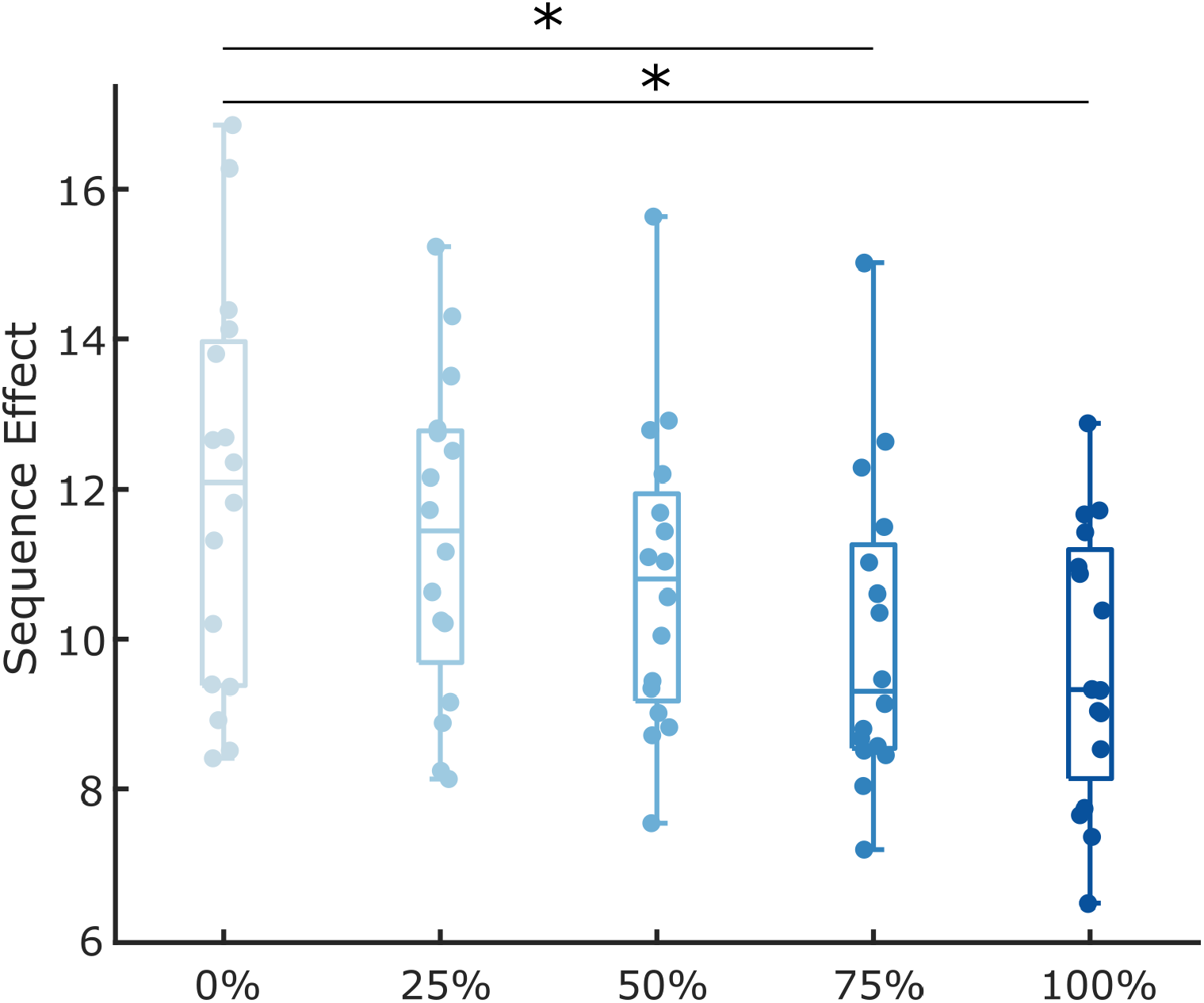
Boxplots comparing the sequence effect across increasing intensities of DBS. Individual data is overlaid. * indicates *p* < 0.05, Bonferroni corrected

### Sequence effect may be further improved by neural closed-loop DBS

The participant displayed the worst sequence effect OFF therapy (Figure 5A), which was improved with clinical olDBS (Fig. 5B). NclDBS further improved the sequence effect from clinical olDBS (Fig. 5C). NclDBS improved the sequence effect in 8 out of the 10 NclDBS conditions across various delay periods between the two hands (Table 2).

**Figure 5.**
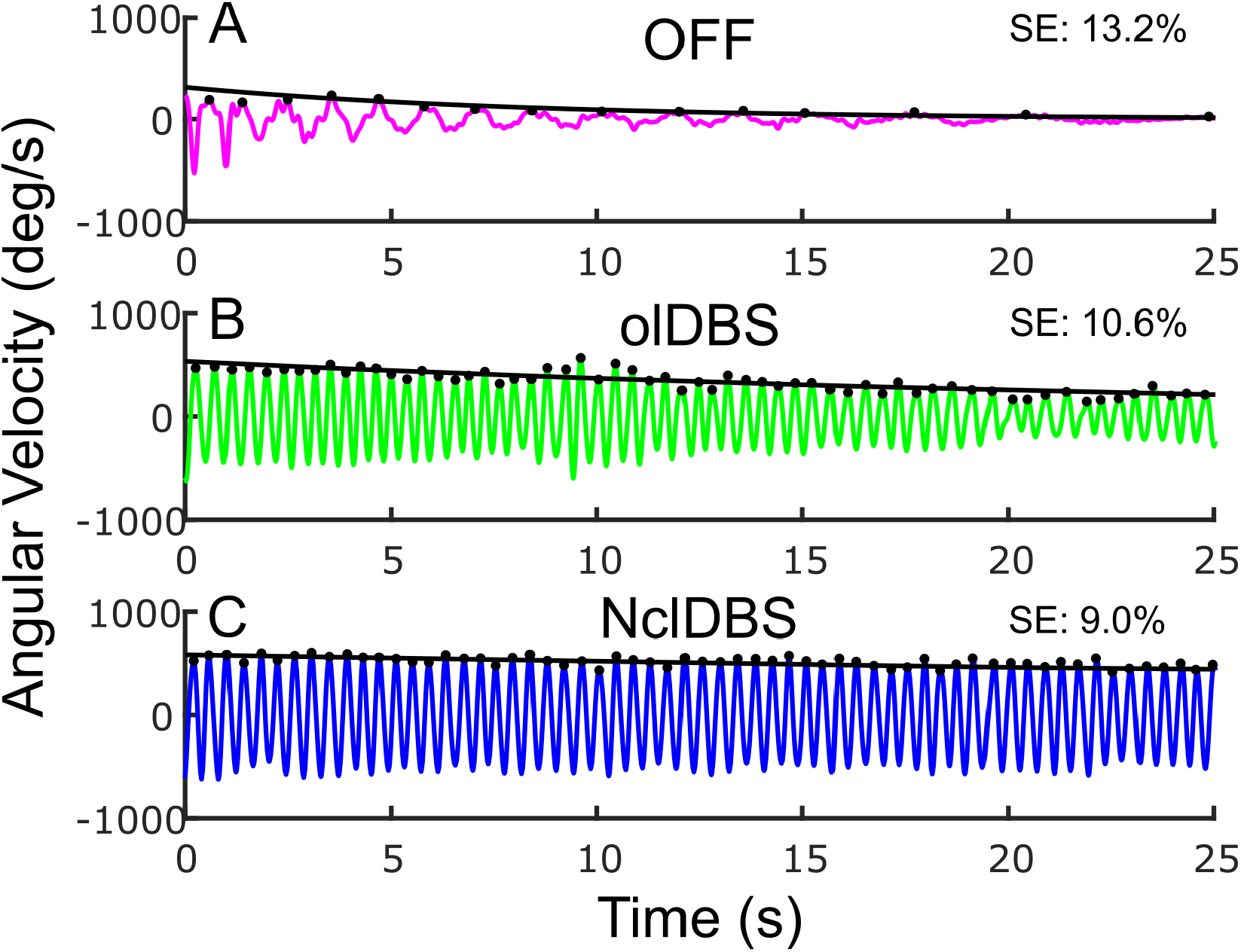
Example of Wrist Flexion Extension (WFE) (A) OFF therapy, (B) open-loop clinical stimulation, (C). neural closed-loop stimulation. Peaks from each WFE cycle indicated by black dots with the fitted exponential curve overlaid.

## Discussion

This study found that an objective normalized metric of the sequence effect in bradykinesia worsened over time in a longitudinal cohort studied off therapy, demonstrating that it was also related to the progression of Parkinson’s disease. The sequence effect improved during STN continuous open-loop DBS in a ‘dose-dependent’ manner and early pilot data suggested that adaptive DBS using a neural controller may provide even better treatment of the sequence effect.

### Measuring the sequence effect in limb movement

One of the difficulties in assessing evidence of the sequence effect and its response to therapies in Parkinson’s disease is the paucity of measurement tools that can differentiate the sequence effect from other metrics such as amplitude and frequency. The MDS-Unified Parkinson’s Disease Rating Scale motor subscale (MDS-UPDRS III) groups together assessments of impairment in amplitude, frequency, and sequence effect into one integer for each item related to limb bradykinesia. Consequently, one cannot discern a specific metric of the sequence effect from the MDS-UPDRS III. Even with quantitative measures, the measure of the sequence effect has varied from a comparison of the first and last multiple of cycles of repetitive movement or gait [33] to linear regressions of time series data [7,34] [12]. We determined that an exponential fit best represented the sequence effect in time series data for instrumented repetitive wrist flexion-extension. The exponential curve best representing sequence effect was personalized to each participant’s wrist flexion extension data by an automated algorithm. The automated algorithm utilized our aforementioned method (i.e., monitoring percent change of moving average both respective to the subsequent peak as well as the next two peaks) to fit an exponential decay or growth function to epochs of sequence effect demonstrated in the angular velocity trace. The majority of trials (63%) demonstrated only one epoch; for all cases (one or multiple epochs) we designated the first epoch as the outcome variable for sequence effect behavior. It will be interesting to further investigate how the sequence effect of multiple epochs varies during self-paced repetitive movement and whether this has disease-related significance, but this was not the focus of the study.

### Sequence effect is a feature of Parkinson’s disease

In this study the sequence effect, or the steepness of decay of the first epoch during movement, was highly correlated with overall PD motor disability (MDS-UPDRS III) and this was not solely due to lower overall velocities in later stages of disease, as it was normalized by initial peak velocity. This suggests that the sequence effect in a simple, short, instrumented upper extremity task could be useful as a surrogate for overall disease severity in remote care and in clinical trials.

The sequence effect was also highly correlated with the bradykinesia sub-score on the MDS-UPDRS III; this is expected as it is defined as “slowness of movement and decrement in amplitude or speed (sequence effect) as movements are continued”, which is similar to its definition in the first formal diagnostic criteria for PD [13,35]. Notably, in this study the sequence effect explained about 35% of the overall MDS-UPDRS III bradykinesia integer sub-score, which includes other components such as the amplitude and speed of the movement. Quantitative, normalized measures of the sequence effect therefore allow more specific investigations into its underlying mechanism and the effects of therapy.

The sequence effect appears to be specific to PD and Multiple System Atrophy (MSA) but does not appear to be a major feature of Progressive Supranuclear Palsy (PSP), Huntington’s disease or dystonia. [1–3,34]. The sequence effect in finger tapping movements has been suggested as a tool to differentiate PSP from PD [3]. It has not been found to correlate with peripheral fatigue or mood [1,22]. The sequence effect in limb bradykinesia has been shown to be a feature of early-stage PD in sequential arm movements, in the Purdue-Pegboard Test, and in finger tapping [4,6,7,34]. In this study, the sequence effect was evident across a broad spectrum of disease severity, even in participants of later stages of PD who present more severe bradykinesia (total MDS-UPDRS III scores>50).

### Sequence effect improved on open and closed-loop STN DBS

One of the debilitating features of the sequence effect in PD is that it does not improve on levodopa or during repetitive transmagnetic stimulation (rTMS)[1,3,20–22,36,37]. Few studies have measured the sequence effect during DBS, although it is well established that DBS improves the overall assessment of bradykinesia from the MDS-UPDRS III. In this study we used randomized presentations of increasing intensities of STN continuous open-loop (ol)DBS, using a single monopole, and found a dose-dependent improvement in the sequence effect in limb bradykinesia during STN olDBS, which was significant at 75% and 100% of the matched clinical DBS intensity. In one subject NclDBS further improved the sequence effect from clinical olDBS in 8 out of the 10 NclDBS conditions providing early evidence that NaDBS may be superior to cDBS for the sequence effect. These results are similar to a case report we previously published, which demonstrated the superiority of an hour of fully embedded NaDBS (Activa™ PC+S-NexusE, Medtronic PLC) for the sequence effect in limb bradykinesia compared to olDBS [24]. Future work can look into evaluating whether the improvement in the sequence effect from NclDBS is more evident over long-term NclDBS and whether this will manifest as a clinically meaningful improvement to people with PD.

### Neural basis of the sequence effect and potential mechanism for the therapeutic effect of DBS

Several studies suggest that the sequence effect may arise from pathological neural activity in cortical, subcortical and cerebellar circuitry that contribute to central drive, motor sequencing, sensorimotor integration, timing cues and updating of the motor set [1,5,7,22,38–42]. Deficits in sensorimotor integration in PD are proposed to be related to irregular and bursting neuronal firing in basal ganglia, pre-motor and supplementary motor circuitry [43–45]. Irregular and bursting neuronal firing patterns increase on medication, which may further impair sensorimotor integration and may contribute to the lack of improvement in the sequence effect from medication [46,47]. Recently, Lofredi et al demonstrated that the duration of low beta (13-20 Hz) bursts in the pathological range predicted the degree of the sequence effect in limb movement [33]. We demonstrated that STN DBS shortened resting state beta band burst durations in a dose-dependent manner [48] and that both 60 Hz and 140 Hz DBS shortened prolonged beta burst durations in people with PD and FOG, while improving their FOG [49]. This suggests that one mechanism for the therapeutic effect of DBS on the sequence effect may be in its ability to attenuate pathological beta burst durations. However, medication may also shorten beta burst durations and medication does not improve the sequence effect, so more investigation into other mechanisms are needed.

### Limitations

The experiments were all performed off medication and thus we cannot assert whether or not medication improved the sequence effect of this particular task, calculated by this metric. This study was focused on the novel metric of the sequence effect, its validation as a measure of bradykinesia and overall PD disease severity, and the effect of DBS. The correlations with bradykinesia and MDS-UPDRS III were performed before activation of DBS. To answer whether the sequence effect became worse over time and to examine the effect of DBS on the sequence effect, testing began after withdrawal of DBS for at least 60 minutes. After turning off STN DBS we have demonstrated that the resting state local field potential spectrum, and specifically beta band power, was stable and unchanged among recordings performed immediately (14 seconds) and every 15 minutes up to 60 minutes later, suggesting that this is enough time to wash out the effect of DBS on underlying pathophysiology [50]. Behavioral studies also suggest that the therapeutic effect of DBS on bradykinesia has lasted well after 60 minutes [51,52]. Any residual effect of DBS on bradykinesia would have biased the data to show less progression compared to baseline and decreased effect of DBS, further supporting these findings. There also may be a ‘cumulative” effect of increasing doses of DBS on behavior as seen for attenuation of the beta band [30]; to avoid this, we presented varying intensities of DBS in a randomized manner, including an OFF DBS condition.

## Conclusions

In this study we demonstrated that the sequence effect worsened over time off therapy over three years. In a careful examination of randomized presentations of different intensities of STN DBS, we showed for the first time that open-loop (continuous) DBS improved the sequence effect of limb bradykinesia in a dose-dependent manner. Finally, we presented early evidence that closed-loop, or adaptive, DBS also improved the sequence effect and was slightly more efficacious than open-loop DBS. This suggests that STN DBS is a promising therapy to improve the sequence effect in bradykinesia for people with Parkinson’s disease.

## Data Availability

All data produced in the present study are available upon reasonable request to the authors

## Author Contributions

Yasmine Kehnemouyi: conceptualization, methodology, software, validation, formal analysis, investigation, data curation, writing – original draft, writing – review & editing, visualization, project administration; Matthew Petrucci: conceptualization, methodology, software, validation, formal analysis, investigation, data curation, writing – original draft, writing – review & editing, visualization, project administration; Kevin Wilkins: conceptualization, methodology, software, validation, formal analysis, investigation, data curation, writing – original draft, writing – review & editing, visualization, project administration; Helen Bronte-Stewart: conceptualization, methodology, investigation, writing – original draft, writing – review & editing, supervision, project administration, funding acquisition

## Acknowledgments

We would like to thank Chioma Anidi and Muhammad Furqan Afzal for their contributions to experimental design and data collection for the study. We would also like to thank the rest of the members of the Human Motor Control and Neuromodulation laboratory, Dr. Jaimie Henderson, and, most importantly, the participants who dedicated their time to this study.

## Funding

This study was funded by the NIH Brain Initiative Grant 1UH3NS107709, NINDS Grant 5 R21 NS096398-02, PF-FBS-2024 from the Parkinson’s Foundation, Michael J. Fox Foundation (9605), Robert and Ruth Halperin Foundation, John A. Blume Foundation, and Helen M. Cahill Award. Medtronic PLC provided devices but no financial support.

## Competing Interests

H.M.B.-S. serves on a clinical advisory board for Medtronic PLC.

## Notes

### Competing Interest Statement

Helen Bronte-Stewart serves on a clinical advisory board for Medtronic PLC.

### Clinical Trial

NCT02384421, NCT01990313, NCT04043403

### Author Declarations

IRB of Stanford University gave ethical approval for this work

## References

[1] Stegemöller EL, Allen DP, Simuni T, MacKinnon CD. Rate-dependent impairments in repetitive finger movements in patients with Parkinson’s disease are not due to peripheral fatigue. Neuroscience Letters 2010;482:1–6. https://doi.org/10.1016/j.neulet.2010.06.054.

[2] Agostino R, Berardelli A, Formica A, Accornero N, Manfredi M. SEQUENTIAL ARM MOVEMENTS IN PATIENTS WITH PARKINSON’S DISEASE, HUNTINGTON’S DISEASE AND DYSTONIA. Brain 1992;115:1481–95. https://doi.org/10.1093/brain/115.5.1481.

[3] Ling H, Massey LA, Lees AJ, Brown P, Day BL. Hypokinesia without decrement distinguishes progressive supranuclear palsy from Parkinson’s disease. Brain 2012;135:1141–53. https://doi.org/10.1093/brain/aws038.

[4] Kang SY, Wasaka T, Shamim EA, Auh S, Ueki Y, Dang N, et al. The Sequence Effect in De Novo Parkinson’s Disease. JMD 2011;4:38–40. https://doi.org/10.14802/jmd.11006.

[5] Railo H, Nokelainen N, Savolainen S, Kaasinen V. Deficits in monitoring self-produced speech in Parkinson’s disease. Clinical Neurophysiology 2020;131:2140–7. https://doi.org/10.1016/j.clinph.2020.05.038.

[6] Koop MM, Shivitz N, Brontë-Stewart H. Quantitative measures of fine motor, limb, and postural bradykinesia in very early stage, untreated Parkinson’s disease. Mov Disord 2008;23:1262–8. https://doi.org/10.1002/mds.22077.

[7] Lee E, Lee JE, Yoo K, Hong JY, Oh J, Sunwoo MK, et al. Neural correlates of progressive reduction of bradykinesia in de novo Parkinson’s disease. Parkinsonism & Related Disorders 2014;20:1376–81. https://doi.org/10.1016/j.parkreldis.2014.09.027.

[8] Nieuwboer A, Dom R, De Weerdt W, Desloovere K, Fieuws S, Broens-Kaucsik E. Abnormalities of the spatiotemporal characteristics of gait at the onset of freezing in Parkinson’s disease. Mov Disord 2001;16:1066–75. https://doi.org/10.1002/mds.1206.

[9] Iansek R, Huxham F, McGinley J. The sequence effect and gait festination in Parkinson disease: Contributors to freezing of gait? Mov Disord 2006;21:1419–24. https://doi.org/10.1002/mds.20998.

[10] Iansek R, Danoudis M. Freezing of Gait in Parkinson’s Disease: Its Pathophysiology and Pragmatic Approaches to Management. Mov Disord Clin Pract 2017;4:290–7. https://doi.org/10.1002/mdc3.12463.

[11] Chee R, Murphy A, Danoudis M, Georgiou-Karistianis N, Iansek R. Gait freezing in Parkinson’s disease and the stride length sequence effect interaction. Brain 2009;132:2151– 60. https://doi.org/10.1093/brain/awp053.

[12] Fasano A, Schlenstedt C, Herzog J, Plotnik M, Rose FEM, Volkmann J, et al. Split-belt locomotion in Parkinson’s disease links asymmetry, dyscoordination and sequence effect. Gait & Posture 2016;48:6–12. https://doi.org/10.1016/j.gaitpost.2016.04.020.

[13] Gibb WR, Lees AJ. The relevance of the Lewy body to the pathogenesis of idiopathic Parkinson’s disease. Journal of Neurology, Neurosurgery & Psychiatry 1988;51:745–52. https://doi.org/10.1136/jnnp.51.6.745.

[14] Agostino R, Berardelli A, Formica A, Stocchi F, Accornero N, Manfredi M. Analysis of repetitive and nonrepetitive sequential arm movements in patients with Parkinson’s disease. Mov Disord 1994;9:311–4. https://doi.org/10.1002/mds.870090305.

[15] Espay AJ, Giuffrida JP, Chen R, Payne M, Mazzella F, Dunn E, et al. Differential response of speed, amplitude, and rhythm to dopaminergic medications in Parkinson’s disease. Mov Disord 2011;26:2504–8. https://doi.org/10.1002/mds.23893.

[16] Hasan H, Burrows M, Athauda DS, Hellman B, James B, Warner T, et al. The BRadykinesia Akinesia INcoordination (BRAIN) Tap Test: Capturing the Sequence Effectin. Mov Disord Clin Pract 2019;6:462–9. https://doi.org/10.1002/mdc3.12798.

[17] Zham P, Kumar DK, Dabnichki P, Poosapadi Arjunan S, Raghav S. Distinguishing Different Stages of Parkinson’s Disease Using Composite Index of Speed and Pen-Pressure of Sketching a Spiral. Front Neurol 2017;8:435. https://doi.org/10.3389/fneur.2017.00435.

[18] Bologna M, Paparella G, Fasano A, Hallett M, Berardelli A. Evolving concepts on bradykinesia. Brain 2020;143:727–50. https://doi.org/10.1093/brain/awz344.

[19] Skodda S, Schlegel U. Speech rate and rhythm in Parkinson’s disease: Speech Rate and Rhythm in Parkinson’s Disease. Mov Disord 2008;23:985–92. https://doi.org/10.1002/mds.21996.

[20] Fabbri M, Guimarães I, Cardoso R, Coelho M, Guedes LC, Rosa MM, et al. Speech and Voice Response to a Levodopa Challenge in Late-Stage Parkinson’s Disease. Front Neurol 2017;8:432. https://doi.org/10.3389/fneur.2017.00432.

[21] Zham P, Poosapadi SA, Kempster P, Raghav S, Nagao KJ, Wong K, et al. Differences in Levodopa Response for Progressive and Non-Progressive Micrographia in Parkinson’s Disease. Front Neurol 2021;12:665112. https://doi.org/10.3389/fneur.2021.665112.

[22] Tinaz S, Pillai AS, Hallett M. Sequence Effect in Parkinson’s Disease Is Related to Motor Energetic Cost. Front Neurol 2016;7. https://doi.org/10.3389/fneur.2016.00083.

[23] Bologna M, Fasano A, Modugno N, Fabbrini G, Berardelli A. Effects of subthalamic nucleus deep brain stimulation and L-DOPA on blinking in Parkinson’s disease. Experimental Neurology 2012;235:265–72.

[24] Bronte-Stewart HM, Petrucci MN, O’Day JJ, Afzal MF, Parker JE, Kehnemouyi YM, et al. Perspective: Evolution of Control Variables and Policies for Closed-Loop Deep Brain Stimulation for Parkinson’s Disease Using Bidirectional Deep-Brain-Computer Interfaces. Front Hum Neurosci 2020;14:353. https://doi.org/10.3389/fnhum.2020.00353.

[25] Petrucci MN, Neuville RS, Afzal MF, Velisar A, Anidi CM, Anderson RW, et al. Neural closed-loop deep brain stimulation for freezing of gait. Brain Stimulation 2020;13:1320–2. https://doi.org/10.1016/j.brs.2020.06.018.

[26] Brontë-Stewart H, Louie S, Batya S, Henderson JM. Clinical motor outcome of bilateral subthalamic nucleus deep-brain stimulation for Parkinson’s disease using image-guided frameless stereotaxy. Neurosurgery 2010. https://doi.org/10.1227/NEU.0b013e3181ecc887.

[27] Quinn EJ, Blumenfeld Z, Velisar A, Koop MM, Shreve LA, Trager MH, et al. Beta oscillations in freely moving Parkinson’s subjects are attenuated during deep brain stimulation. Movement Disorders 2015. https://doi.org/10.1002/mds.26376.

[28] Koop MM, Andrzejewski A, Hill BC, Heit G, Bronte-Stewart HM. Improvement in a quantitative measure of bradykinesia after microelectrode recording in patients with Parkinson’s disease during deep brain stimulation surgery. Movement Disorders 2006. https://doi.org/10.1002/mds.20796.

[29] Koop MM, Shivitz N, Brontë-Stewart H. Quantitative measures of fine motor, limb, and postural bradykinesia in very stage, untreated parkinson’s disease. Movement Disorders 2008. https://doi.org/10.1002/mds.22077.

[30] Louie S, Koop MM, Frenklach A, Bronte-Stewart H. Quantitative lateralized measures of bradykinesia at different stages of Parkinson’s disease: The role of the less affected side. Movement Disorders 2009. https://doi.org/10.1002/mds.22741.

[31] Blumenfeld Z, Koop MM, Prieto TE, Shreve LA, Velisar A, Quinn EJ, et al. Sixty-hertz stimulation improves bradykinesia and amplifies subthalamic low-frequency oscillations. Movement Disorders : Official Journal of the Movement Disorder Society 2017;32:80–8. https://doi.org/10.1002/mds.26837.

[32] Petrucci MN, Anderson RW, O’Day JJ, Kehnemouyi YM, Herron JA, Bronte-Stewart HM. A Closed-loop Deep Brain Stimulation Approach for Mitigating Burst Durations in People with Parkinson’s Disease. 2020 42nd Annual International Conference of the IEEE Engineering in Medicine & Biology Society (EMBC), Montreal, QC, Canada: IEEE; 2020, p. 3617–20. https://doi.org/10.1109/EMBC44109.2020.9176196.

[33] Lofredi R, Tan H, Neumann W-J, Yeh C-H, Schneider G-H, Kühn AA, et al. Beta bursts during continuous movements accompany the velocity decrement in Parkinson’s disease patients. Neurobiology of Disease 2019;127:462–71. https://doi.org/10.1016/j.nbd.2019.03.013.

[34] Djurić-Jovičić M, Petrović I, Ječmenica-Lukić M, Radovanović S, Dragašević-Mišković N, Belić M, et al. Finger tapping analysis in patients with Parkinson’s disease and atypical parkinsonism. Journal of Clinical Neuroscience 2016;30:49–55.

[35] Postuma RB, Berg D, Stern M, Poewe W, Olanow CW, Oertel W, et al. MDS clinical diagnostic criteria for Parkinson’s disease. Mov Disord 2015;30:1591–601. https://doi.org/10.1002/mds.26424.

[36] Kang SY, Wasaka T, Shamim EA, Auh S, Ueki Y, Lopez GJ, et al. Characteristics of the sequence effect in Parkinson’s disease: Sequence Effect in Parkinson’s Disease. Mov Disord 2010;25:2148–55. https://doi.org/10.1002/mds.23251.

[37] Wu T, Zhang J, Hallett M, Feng T, Hou Y, Chan P. Neural correlates underlying micrographia in Parkinson’s disease. Brain 2016;139:144–60. https://doi.org/10.1093/brain/awv319.

[38] Maier MA, Bennett KM, Hepp-Reymond MC, Lemon RN. Contribution of the monkey corticomotoneuronal system to the control of force in precision grip. Journal of Neurophysiology 1993;69:772–85. https://doi.org/10.1152/jn.1993.69.3.772.

[39] Lüders HO. The supplementary sensorimotor area. An overview. Adv Neurol 1996;70:1– 16.

[40] Rodriguez-Oroz MC, Jahanshahi M, Krack P, Litvan I, Macias R, Bezard E, et al. Initial clinical manifestations of Parkinson’s disease: features and pathophysiological mechanisms. Lancet Neurol 2009;8:1128–39. https://doi.org/10.1016/S1474-4422(09)70293-5.

[41] Koop MM, Hill BC, Bronte-Stewart HM. Perceptual errors increase with movement duration and may contribute to hypokinesia in Parkinson’s disease. Neuroscience 2013;243:1–13. https://doi.org/10.1016/j.neuroscience.2013.03.026.

[42] Cao S-S, Yuan X-Z, Wang S-H, Taximaimaiti R, Wang X-P. Transverse Strips Instead of Wearable Laser Lights Alleviate the Sequence Effect Toward a Destination in Parkinson’s Disease Patients With Freezing of Gait. Front Neurol 2020;11:838. https://doi.org/10.3389/fneur.2020.00838.

[43] Moore AP. Impaired sensorimotor integration in parkinsonism and dyskinesia: a role for corollary discharges? Journal of Neurology, Neurosurgery & Psychiatry 1987;50:544–52. https://doi.org/10.1136/jnnp.50.5.544.

[44] Rossini PM, Filippi MM, Vernieri F. Neurophysiology of sensorimotor integration in Parkinson’s disease. Clin Neurosci 1998;5:121–30.

[45] Lewis GN. Altered sensorimotor integration in Parkinson’s disease. Brain 2002;125:2089– 99. https://doi.org/10.1093/brain/awf200.

[46] Filion M, Tremblay L. Abnormal spontaneous activity of globus pallidus neurons in monkeys with MPTP-induced parkinsonism. Brain Research 1991;547:140–4. https://doi.org/10.1016/0006-8993(91)90585-J.

[47] Bronte-Stewart HM. Postural instability in idiopathic Parkinson’s disease: the role of medication and unilateral pallidotomy. Brain 2002;125:2100–14. https://doi.org/10.1093/brain/awf207.

[48] Anderson RW, Kehnemouyi YM, Neuville RS, Wilkins KB, Anidi CM, Petrucci MN, et al. A novel method for calculating beta band burst durations in Parkinson’s disease using a physiological baseline. Journal of Neuroscience Methods 2020;343:108811. https://doi.org/10.1016/j.jneumeth.2020.108811.

[49] Anidi C, O’Day JJ, Anderson RW, Afzal MF, Syrkin-Nikolau J, Velisar A, et al. Neuromodulation targets pathological not physiological beta bursts during gait in Parkinson’s disease. Neurobiology of Disease 2018;120:107–17. https://doi.org/10.1016/j.nbd.2018.09.004.

[50] Trager MH, Koop MM, Velisar A, Blumenfeld Z, Nikolau JS, Quinn EJ, et al. Subthalamic beta oscillations are attenuated after withdrawal of chronic high frequency neurostimulation in Parkinson’s disease. Neurobiology of Disease 2016;96:22–30. https://doi.org/10.1016/j.nbd.2016.08.003.

[51] Temperli P, Ghika J, Villemure J-G, Burkhard PR, Bogousslavsky J, Vingerhoets FJG. How do parkinsonian signs return after discontinuation of subthalamic DBS? Neurology 2003;60:78–81.

[52] Cooper SE, Driesslein KG, Noecker AM, McIntyre CC, Machado AM, Butson CR. Anatomical targets associated with abrupt versus gradual washout of subthalamic deep brain stimulation effects on bradykinesia. PLoS One 2014;9:e99663.

